# Initial protection against Omicron in children and adolescents by BNT162b2

**DOI:** 10.1101/2022.05.22.22275323

**Authors:** Ofra Amir, Yair Goldberg, Micha Mandel, Yinon M. Bar-On, Omri Bodenheimer, Laurence Freedman, Nachman Ash, Sharon Alroy-Preis, Amit Huppert, Ron Milo

## Abstract

**BACKGROUND:** The BNT162b2 (Pfizer-BioNTech) 2-dose vaccine for children and the BNT162b2 3rd dose for adolescents were approved shortly before the Omicron outbreak in Israel. The effects of these vaccines on the rates of Omicron confirmed infection are not yet clear.

**METHODS:** We extracted data for the Omicron-dominated (sub-lineage BA.1) period December 26, 2021 through January 8, 2022. We compared rates of confirmed Covid-19 infection between children 5-10 years old 14-35 days after receiving the 2nd dose to an internal control group of children 3-7 days after receiving the 1st dose (when the vaccine is not yet effective). Similarly, we compared confirmed infection rates in adolescents 12-15 years old 14-60 days after receiving a booster dose to an internal control group of adolescents 3-7 days after receiving the booster dose. We used Poisson regression, adjusting for age, sex, socioeconomic status, calendar week, and exposure.

**RESULTS:** In the 5-10 age group, the estimated rate of confirmed infection was 2.3 fold (95% CI, 2.0 to 2.5) lower in the 2nd dose group than in the internal control group. In adolescents, the third dose decreased confirmed infection rates by 3.3-fold (95% CI, 2.8 to 4.0).

**CONCLUSIONS:** A recent 2-dose BNT162b2 vaccination in children and a recent booster dose in adolescents reduced the rate of confirmed infection compared to the respective internal control groups. Future studies are needed to assess the duration of this protection and protection against other outcomes such as PIMS and long-COVID.

## Introduction

The effectiveness of the BNT162b2 vaccine has been shown to be poorer against the Omicron than against the Delta and other variants.^1^ However, there is little evidence about the real-world effectiveness of the BNT162b2 vaccine against the Omicron variant in children and adolescents. Here we used data from Israel to explore the effectiveness of the BNT162b2 2-dose regimen for children 5-11 years old and the additional booster dose for adolescents 12-15 years old, which were approved shortly before the Omicron outbreak in Israel.

In Israel, the BNT162b2 vaccine was approved for adolescents aged 12-15 on June 2, 2021, and a booster dose was approved starting August 29, 2021, for individuals who received the 2nd dose at least 5 months previously. For children aged 5-11, the 2-dose vaccination (using a third of the dosage given for ages 12 and above) was administered starting November 23, 2021. We examine the short-term protection conferred by the BNT162b2 vaccine against confirmed infection in children (two doses) and adolescents (third dose) from December 26, 2021, to January 8, 2022, a period when the Omicron variant was dominant in Israel.^2^ Due to significant policy changes in testing and isolation of contacts and quarantine in schools, reliable estimates of effectiveness are difficult to obtain for the period after January 8, 2022.

For children, recent studies from the US estimated vaccine effectiveness (VE) of 65% in the first two weeks after vaccination, followed by rapid waning.^3,4^ The VE was lower for children than for the 12-15 age group. A report from the CDC based on a cohort of about 1,000 children aged 5-11 estimated a VE of 31% up to 82 days from the date of vaccination.^5^ In adolescents 16-18 years old, a booster dose was shown to lower the confirmed infection rate by a factor of 3.7 compared to two doses during the Delta wave,^6^ For adolescents aged 12-15, a booster dose was shown to improve protection against Omicron infection by a factor of 2.9 compared to two doses.^4^ In this study, we estimated (1) the adjusted rates of confirmed infection following two doses of the BNT162b2 vaccine in children aged 5-10 up to 35 days from the 2nd dose, and (2) the adjusted rates of confirmed infections following a third dose of the BNT162b2 vaccine in adolescents aged 12-15 up to 60 days from this dose.

## Methods

### STUDY POPULATION

We analyzed observational data on SARS-Cov-2 infections collected during Israel’s fifth wave, which was Omicron-dominant (BA.1 sub-lineage). We used the Israeli Ministry of Health database that includes information on all vaccinations and tests that had been conducted in Israel. The study population included children (age 5-10) and adolescents (12-15) who were either vaccinated or took at least one test (PCR or state-regulated antigen) before December 1st, 2021. We excluded the 11 year-old age cohort, as our data included age in years only, and vaccination eligibility dates differed between 11 and 12 year-olds. Thus, some of the ‘11 year-old group’ turned 12 before or during the study period. We excluded persons who had documented positive PCR or state-regulated antigen results prior to the study period, had stayed abroad during the entire study period, or had received a vaccine different from BNT162b2 before the end of the study period; see Figure 1 for details.

**Figure 1:**
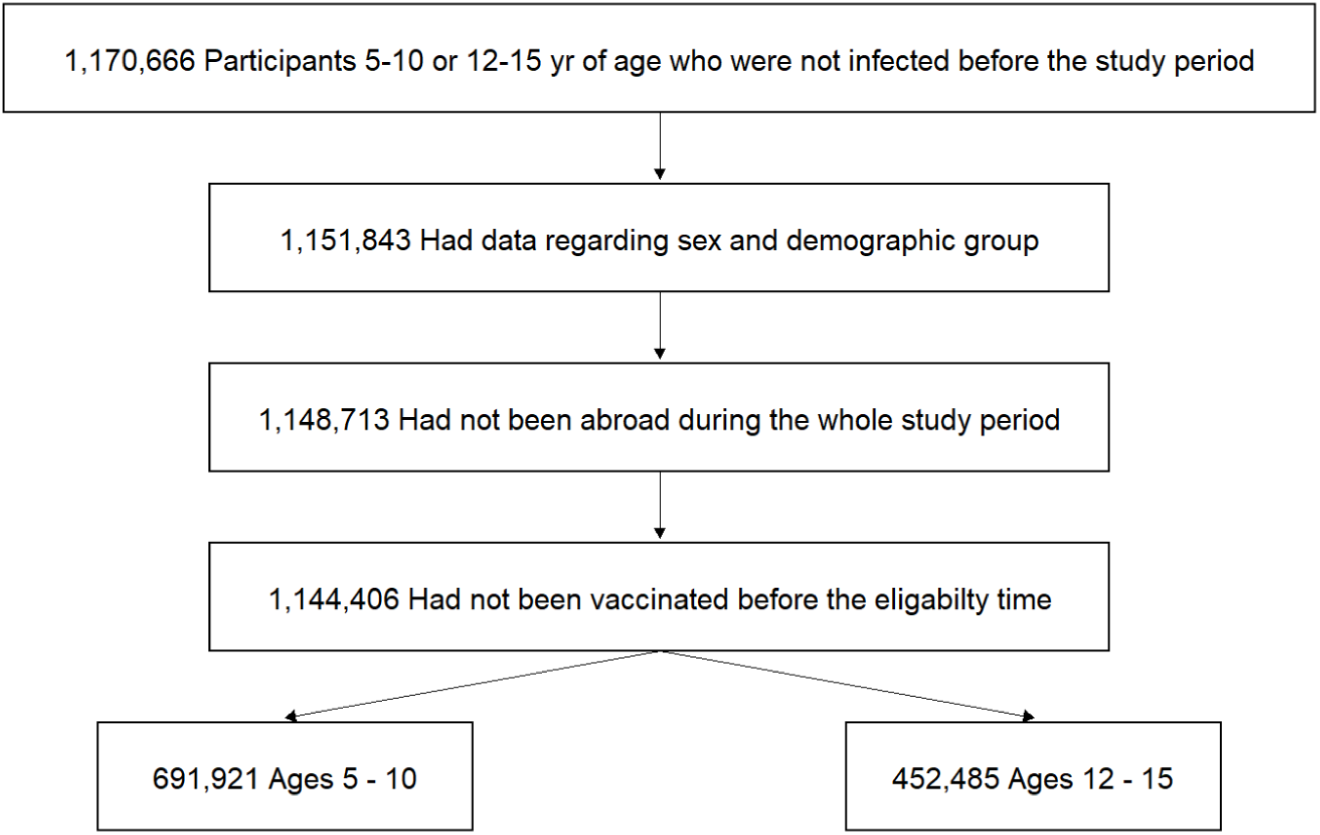
The study population included persons who were between the ages of 5-10 or 12-15, had no documented positive PCR, or a positive state-regulated Antigen result prior to the study period, had not stayed abroad during the whole study period, had not been vaccinated before their age group eligibility time and had not been vaccinated with a vaccine different from BNT162b2 before the beginning of the study period.

### STUDY DESIGN

Our study compared the confirmed infection rates in different cohorts (defined below) during a two-week study period, from December 26, 2021 to January 8, 2022 in which the national surveillance testing was functioning well, and before home antigen testing became common. Children in the 5-10 age group were divided into three cohorts: those unvaccinated, those who received the second dose of vaccine at least 14 days previously, and an internal control cohort of those who received their first dose 3-7 days previously. Adolescents in the 12-15 age group were divided into six cohorts: those unvaccinated, those who received two doses of the vaccine only (divided according to time since the second dose (14-60, 60-120, and 120 or more days), those who received a booster (third) dose at least 14 days previously, and an internal control cohort of those who received the booster dose 3-7 days previously. Cohort membership was dynamic, that is, during the study period an individual could leave one cohort and join another.

Differences in behavioral and exposure characteristics,^7,8^ and possibly in the proportion of undocumented prior infections,^9^ might bias comparisons between the 2-dose and the unvaccinated cohorts in children and between the booster dose cohorts and other cohorts in adolescents. To mitigate such biases, our main analysis focused on comparisons to the internal control cohorts that had received the vaccine, but before the vaccine was expected to affect their risk of confirmed infection.^10^

Since the two age groups became eligible to receive either the 2-dose vaccination or the 3rd dose, respectively, not long before the study period, our study estimated only the short-term effectiveness of these vaccinations. Figure S1 presents the percentages of vaccinees over time. The main analysis of the 12-15 age group focused on assessing the effectiveness of the 3rd dose since a relatively small portion of individuals in this age group received the 2nd dose during the relevant time period. The main analysis included only the General Jewish sector, as vaccination rates were very low in the Arab and Jewish ultra-orthodox sectors.

### STATISTICAL ANALYSIS

We analyzed the data using methodology similar to that used in our previous studies.^11^ The number of confirmed infections and the number of days at risk during the study period were counted for each cohort. For each age group, a separate Poisson regression model was used to estimate the adjusted rate of confirmed infections per 100,000 risk days, adjusting for age (one-year categories), sex, socioeconomic status (low, medium, high), calendar week, and an exposure risk measure. The latter time-dependent variable was calculated for each person on each follow-up day according to the proportion of new confirmed infections during the past seven days in their area of residence; it was then divided into five categories according to quintiles (see Bar-On et al.^11^ for details). A national average risk was imputed to individuals with missing data on residency.

We conducted several sensitivity analyses. To examine the effect of including all population sectors (General Jewish, Jewish ultra-Orthodox, and Arab), we repeated the analysis on the whole population adding the sector as a covariate in the analysis. While our main analysis included only two weeks until January 8, after which state-regulated tests were supplemented by undocumented home tests for vaccinated individuals, a second analysis extended the study period by an additional one week (to January 15, 2022). We also analyzed confirmed infections over a shorter study period, including only the week of January 2 to January 8, 2022. The shorter time period has the benefit of maintaining a similar rate of exposure throughout the study period.

## Results

Table 1 reports basic descriptive statistics for the main cohorts of interest used in the analysis (see details on other cohorts in Tables S1 and S2). The main analysis (Figure 2 and Table 2) shows that, for children 5-10 years old, two doses provided a 2.3-fold (95% CI, 2.0 to 2.5) decrease in confirmed infection rates. The estimated infection rate of the 2nd dose cohort was 109 (95% CI, 101 to 118) infections per 100,000 risk-days compared to 249 (95% CI, 230 to 269) in the corresponding internal control cohort. In adolescents 12-15, the third dose decreased confirmed infection rates by 3.3-fold (95% CI, 2.8 to 4.0). The estimated infection rate of the booster cohort was 76 (95% CI, 65 to 87) per 100,000 risk-days compared to 251 (95% CI, 229 to 276) in the booster internal control cohort. The sensitivity analyses that considered different study periods and population groups yielded similar results (see Table 3).

**Table 1.** Characteristics of the study groups used in the main analysis. The table presents the proportion of person-days at risk instead of the proportion of individuals. Values are presented for the study period - December 26, 2021 to January 8, 2022.

| Ages 5-10<br>2nd dose effect |  |  |  |  | Ages 12-15<br>3rd dose effect |  |  |  |
| --- | --- | --- | --- | --- | --- | --- | --- | --- |
| Group | 2nd dose internal control<br>(3-7 days from 1st dose)<br>Person-days at risk =<br>367,168 |  | 2nd dose<br>(14-35 days from 2nd<br>dose)<br>Person-days at risk =<br>366,364 |  | 3rd dose internal control<br>(3-7 days from 3rd dose)<br>Person-days at risk =<br>190,139 |  | 3rd dose<br>(14-60 days from 3rd dose)<br>Person-days at risk =<br>178,780 |  |
|  | % person<br>days at risk | # infections | % person<br>days at risk | # infections | % person<br>days at risk | # infections | % person<br>days at risk | # infections |
| Female | 48.5% | 411 | 48.3% | 287 | 48.4% | 265 | 48.9% | 99 |
| Male | 51.5% | 411 | 51.7% | 315 | 51.6% | 253 | 51.1% | 80 |
| General<br>Jewish | 86.7% | 743 | 94.9% | 576 | 94.7% | 494 | 95.8% | 166 |
| Ultra-Orthodox | 6.6% | 69 | 3.2% | 21 | 3.1% | 20 | 2.7% | 13 |
| Arabs | 6.7% | 10 | 1.8% | 5 | 2.1% | 4 | 1.5% | 0 |

**Table 2.** Adjusted rate ratios of study cohorts vs. the vaccinated cohorts of interest (2nd dose in ages 5-10, 3rd dose in ages 12-15) adjusted for age category, gender, socioeconomic status (Low, Medium, High), calendar week, and an exposure risk measure, for the study period - December 26, 2021 to January 8, 2022.

| **Ages 5-10**<br>**2nd dose** |  |  |
| --- | --- | --- |
| Cohort | Confirmed infections<br>(at-risk days) | Adjusted rate ratio vs. 2nd dose |
| Unvaccinated | 10,048<br>(4,420,027) | 2.4 [2.2, 2.6] |
| Internal control | 743 (318,513) | 2.3 [2.0, 2.5] |
| 2nd dose<br>(14-35 days) | 576 (347,726) | Ref |

| **Ages 12-15**<br>**3rd dose effect** |  |  |
| --- | --- | --- |
| Cohort | Confirmed infections<br>(at-risk days) | Adjusted rate ratio vs. 3rd dose |
| Unvaccinated | 2,684 (834,149) | 5.0 [4.3, 5.9] |
| 2nd dose<br>(14-60 days) | 153 (115,371) | 2.2 [1.8, 2.8] |
| 2nd dose<br>(60-120 days) | 1,999 (815,036) | 3.8 [3.3, 4.5] |
| 2nd dose<br>(120+ days) | 5,983<br>(2,003,011) | 4.2 [3.6, 4.9] |
| Internal control | 494 (180,100) | 3.3 [2.8, 4.0] |
| 3rd dose<br>(14-60 days) | 166 (171,281) | Ref |

**Table 3.** Results from sensitivity analyses - inclusion of all population groups (General Jewish, ultra-Orthodox, Arab), longer (December 26, 2021 to January 15, 2022) and shorter (January 2nd, 2022 to January 8th, 2022) study periods.

| Sensitivity analysis | Ages 5-10<br>2nd dose effect |  |  | Ages 12-15<br>3rd dose effect |  |  |
| --- | --- | --- | --- | --- | --- | --- |
|  | 2nd dose confirmed infections (at-risk days) | internal control confirmed infections (at-risk days) | Adjusted rate ratio internal control vs. 2nd dose | 3rd dose confirmed infections (at-risk days) | internal control confirmed infections (at-risk days) | Adjusted rate ratio internal control vs. 3rd dose |
| Main Analysis | 576 (347,726) | 743 (318,513) | 2.3 [2.0, 2.5] | 166 (171,281) | 494 (180,100) | 3.3 [2.8, 4.0] |
| All population groups | 602 (366,364) | 822 (367,168) | 2.3 [2.1, 2.6] | 179 (178,780) | 518 (190,139) | 3.2 [2.7, 3.8] |
| Longer study period | 3,142 (840,479) | 1,699 (430,094) | 1.9 [1.8, 2.0] | 911 (410,853) | 1,379 (285,267) | 2.9 [2.6, 3.1] |
| Shorter study period | 530 (286,144) | 524 (148,127) | 2.2 [2.0, 2.5] | 148 (120,433) | 395 (103,922) | 3.2 [2.7, 3.9] |

**Figure 2:**
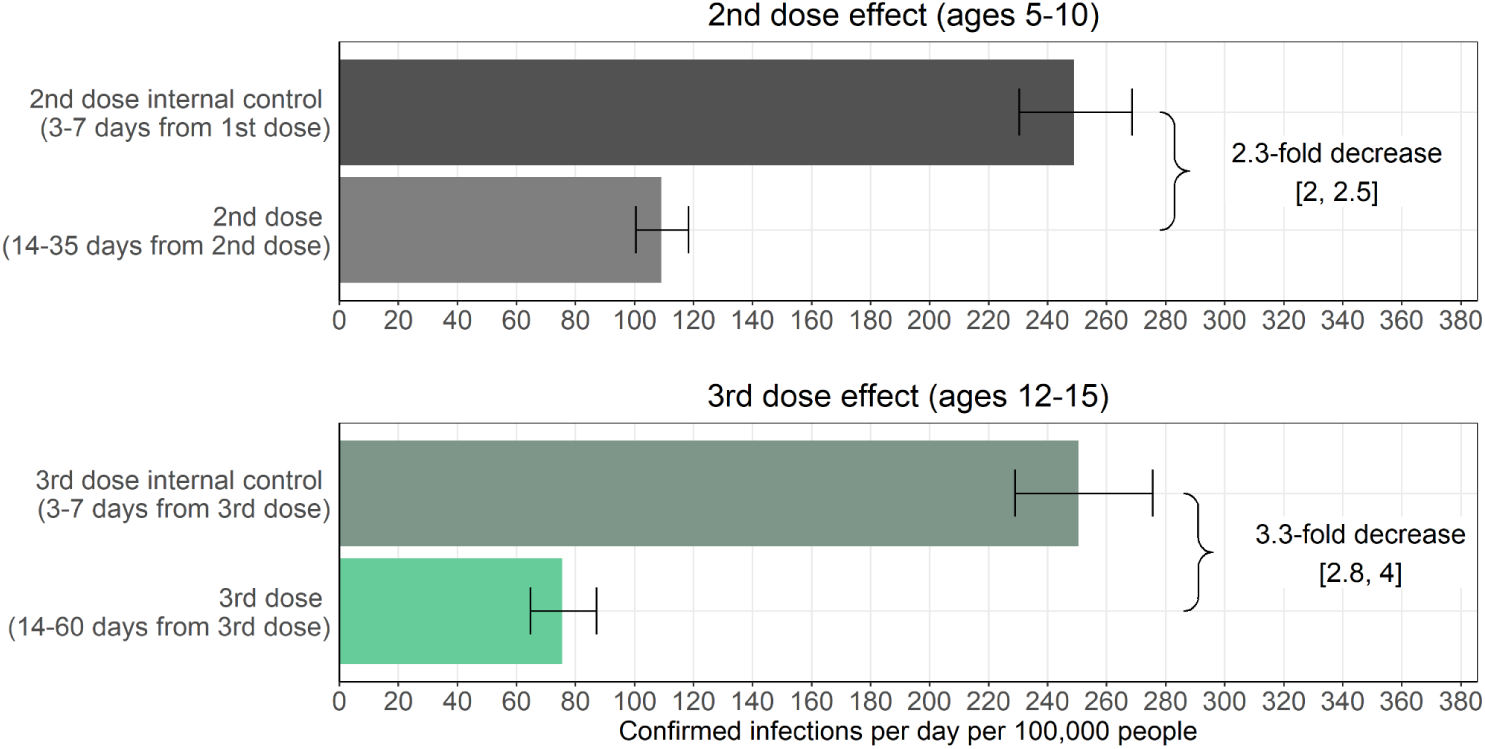
Adjusted rate of confirmed infections per 100,000 risk days obtained from a Poisson regression analysis for the study period December 26, 2021, to January 8, 2022, adjusted for age category, gender, socioeconomic status (Low, Medium, High), calendar week, and exposure. The 95% confidence intervals are not adjusted for multiplicity.

The results of analyses that included unvaccinated children and adolescents who received only two doses are shown in Figure 3 and summarized in Table 2. In the 5-10 age group, the estimated level of protection provided by the 2nd dose was similar when compared to either the internal control cohort or the unvaccinated cohort. In the 12-15 age group, the protection from infection of the 2-dose vaccine waned quickly over time, with infection rates rising from 133 per 100,000 risk days among those who received their second dose between 14 and 60 days previously to 299 among those receiving it more than 120 days previously. Protection was restored and substantially increased with receipt of the 3rd dose. Comparing the 3rd dose to individuals who received the 2nd dose more than 120 days previously yielded a slightly higher protection (4.1-fold lower rate) than the comparison with the internal control group (3.4-fold lower rate).

**Figure 3.**
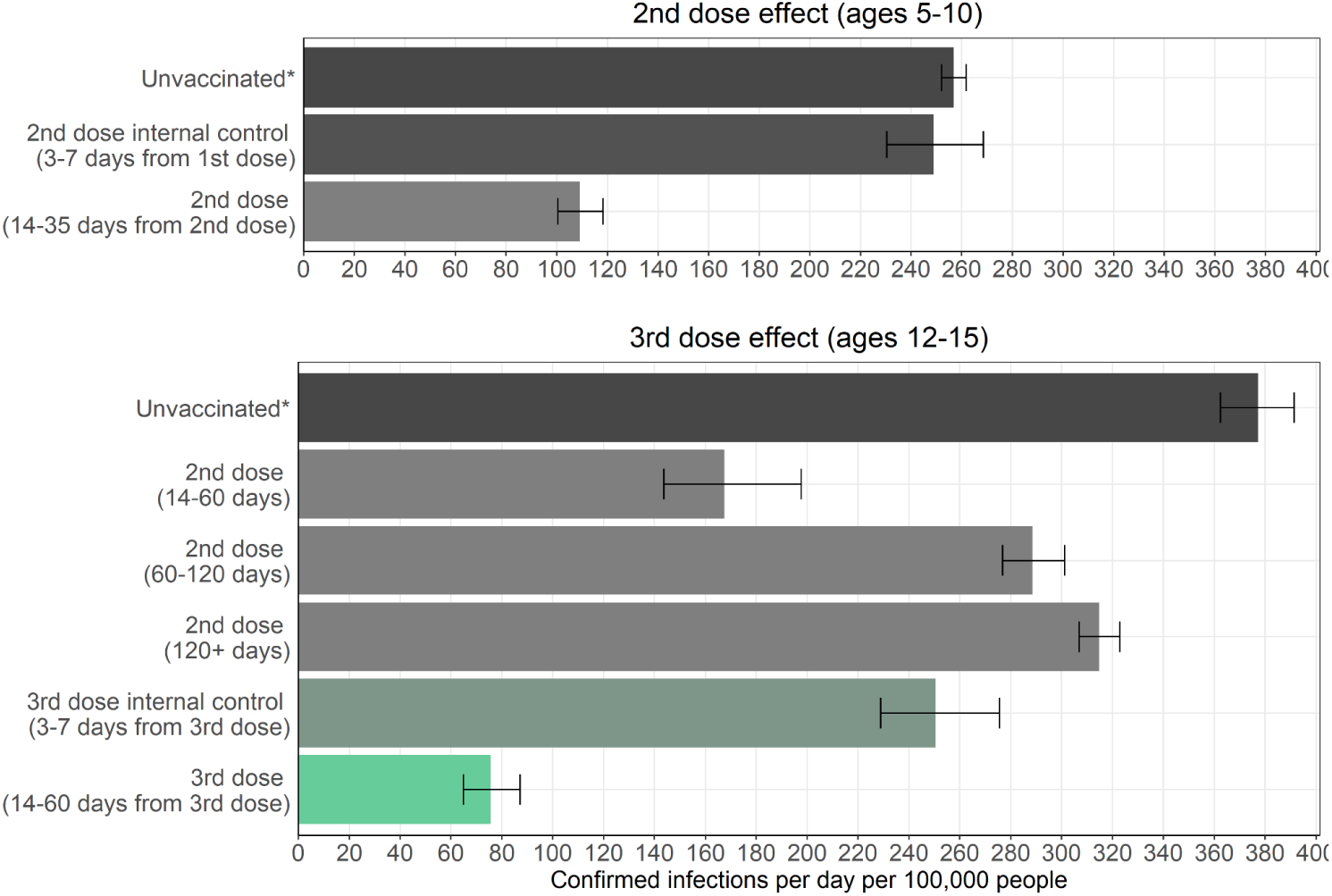
Adjusted rate of confirmed infections per 100,000 risk days obtained from a Poisson regression analysis for the study period December 26, 2021, to January 8, 2022, stratified by age groups and adjusted for age category, gender, socioeconomic status (Low, Medium, High), calendar week, and an exposure risk measure. The 95% confidence intervals are not adjusted for multiplicity. *All groups, and in particular the unvaccinated group, likely have high fractions of individuals with undocumented prior infections. Therefore, the adjusted confirmed infection rates are likely underestimated.

## Discussion

Our results show that the BNT162b2 vaccine provided an initial ≈2-fold increased protection against infection in children 5-10 years old. The estimated protection is in line with the VE results estimated in the US for a similar study period and time from the vaccine.^3,4^ It is somewhat higher than that reported by the CDC,^5^ possibly due to waning immunity, since more time had passed since vaccination in the CDC study. Our analysis further shows that a recent booster dose in adolescents decreased infections by ≈3-4 fold compared to the internal control, which is similar to estimates from the US.^4^

We note that the lower confirmed infection rates in the vaccinated cohorts compared to the unvaccinated are not explained by different testing behavior (see Figure S2). Specifically, individuals in the unvaccinated cohorts in both age groups tested less than those in the vaccinated cohorts, suggesting that the estimated protection compared to unvaccinated individuals might be underestimated. While the unvaccinated cohorts had lower testing rates than the vaccinated cohorts, in the 5-10 age group, the internal control group had a somewhat higher testing rate than the 2nd dose cohort, which may lead to an overestimation of the protection conferred by the vaccine. In the vaccinated 12-15 age group, the internal control group had a lower testing rate than the booster cohort, which may suggest a higher level of protection conferred by the booster than that estimated in our analysis.

This study provides an estimate for the short-term protection against confirmed infection conferred by the BNT162b2 vaccine. Extending the study period and obtaining reliable estimates for a longer follow-up time was not possible for the following reasons. First, starting January 6, 2022, there were changes to the testing and isolation policies in Israel. These changes, which included the extended use of home testing, make it difficult to estimate the rate of confirmed infection in children and adolescents. Second, using the internal control cohorts in later time periods may lead to biases since vaccine rates declined. Moreover, individuals who enter these internal control cohorts after the study period may have higher rates of undocumented prior infections compared to those who vaccinated earlier due to the high exposure during the Omicron wave.

While the current study estimated protection against confirmed infection, the effect of the vaccine on other outcomes in these age groups remains unclear. In particular, estimation of the protection against pediatric inflammatory multisystem syndrome temporally associated with SARS-CoV-2 (PIMS-TS).^12^ and long-COVID, as well as side effects can provide additional important information for policy-making regarding vaccination in these age groups.

## Data Availability

The individual-level data used in this study cannot be publicly shared even if anonymized due to privacy restrictions.

## Supplementary Appendix

### Study Information

#### Ethics statement

The study was approved by the Institutional Review Board of the Sheba Medical Center. Helsinki approval number: SMC-8228-21.

### Supplementary Methods - Database

The analysis is based on the Israel Ministry of Health’s database, as described in our previous studies.^10^ Israel began a vaccination campaign with the BNT162b2 vaccine on December 20, 2020, initially to people aged 60 or older and gradually to younger populations. Starting June 2, 2021, adolescents aged 12-15 were offered the vaccine. Following the FDA approval for children vaccination, children 5-11 year old were eligible to vaccinate starting November 23, 2021. Individuals 12-15 were later allowed to receive a third booster dose starting August 29, 2021, if 5 months passed since the complete 2-dose vaccination. Later the threshold was reduced to 3 months following the complete 2-dose vaccination.

Israel has a centralized health system of four health maintenance organizations (HMOs), where each resident can choose to enroll in one of them. Polymerase Chain Reaction (PCR) tests and Institutional Antigen tests for SARS-CoV-2 infections as well as vaccination against the virus are provided free of charge in designated centers and their results are directly reported to the Ministry of Health (MoH). The MoH established a centralized Covid-19 national database containing regularly updated information on all PCR and state-regulated Antigen tests and results, vaccination dates, and follow-up data on all infected individuals, including the severity of disease and mortality. The MoH database also includes basic demographic information, such as sex, age, place of residency, and population sector.

**Figure S1.**
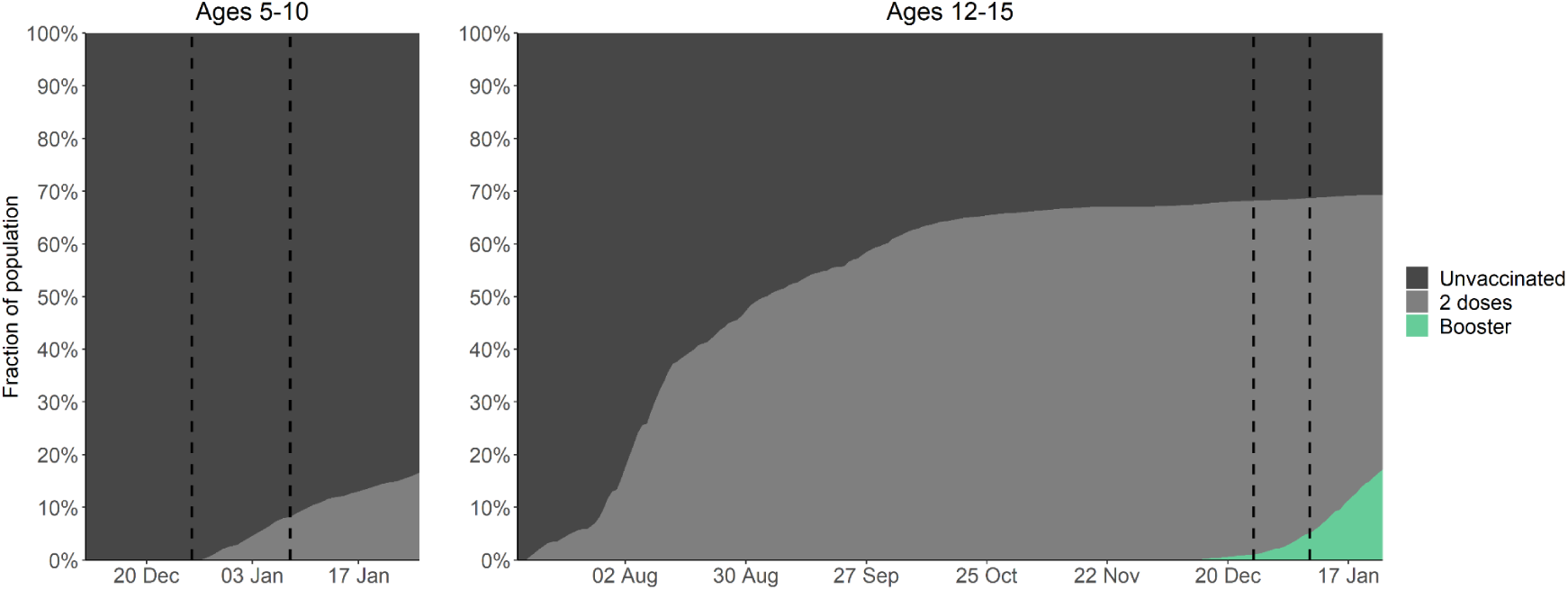
Vaccination rates in the study populations. Vaccination status (percentages) in the 5-10 and 12-15 age groups since becoming eligible to receive the vaccine (June 2, 2021 for the 12-15 age group and November 23, 2021 for the 5-10 age group), and the booster dose (Aug 29, 2021). Vertical dashed lines represent the study period.

**Figure S2.**
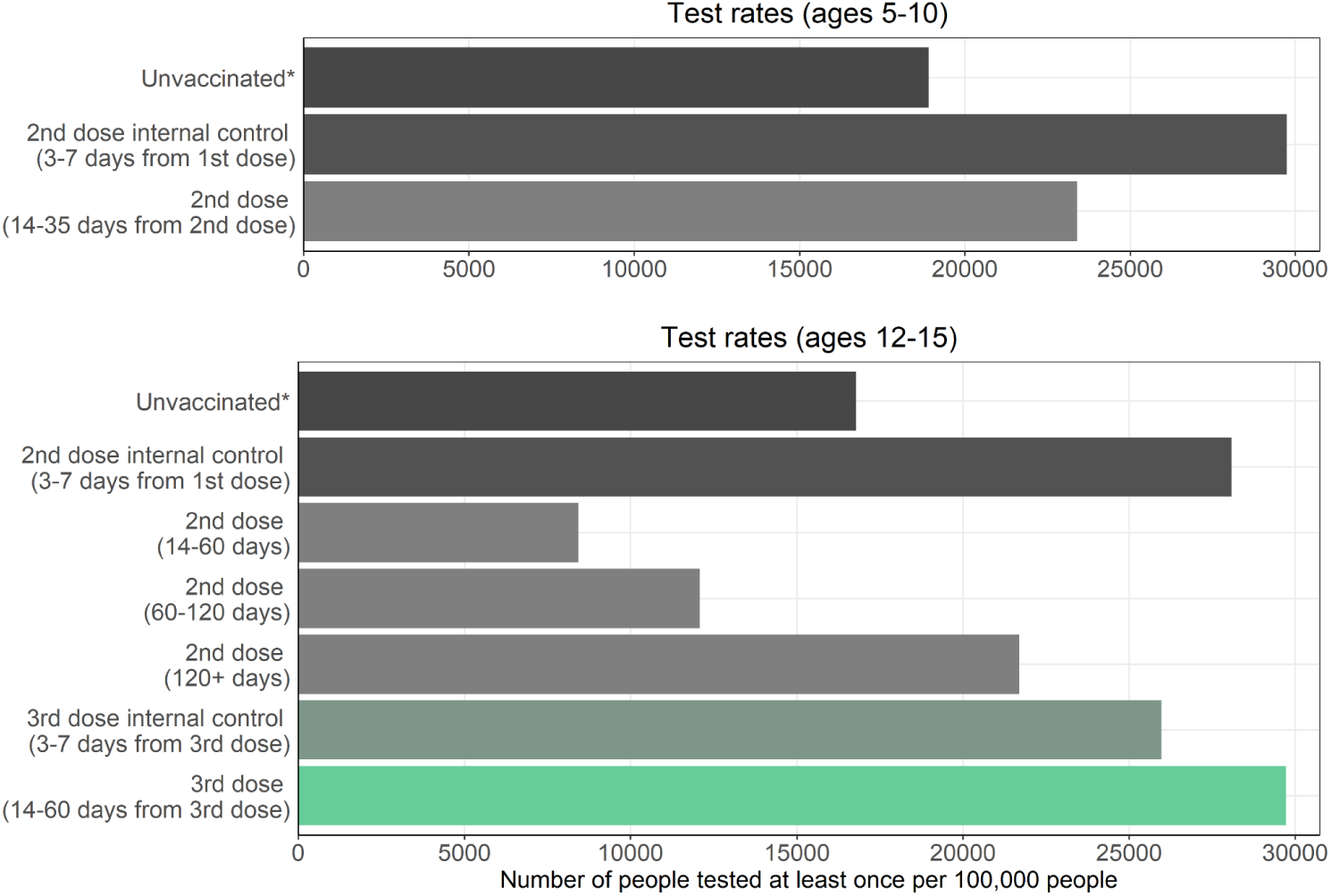
Testing rates in the study populations. The number of people who tested at least once in each epidemiological week per 100,000 people in the different cohorts, during the study period - December 26, 2021 and January 8, 2022.

**Table S1.** Characteristics of the 5-10 age group study cohorts used in the analysis. The table presents the proportion of person-days at risk instead of the proportion of individuals. Values are presented for the study period - December 26, 2021 to January 8, 2022.

| Ages 5-10<br>2nd dose effect |  |  |  |  |  |  |
| --- | --- | --- | --- | --- | --- | --- |
| Group | Unvaccinated* |  | 2nd dose internal control<br>(3-7 days from 1st dose) |  | 2nd dose<br>(14-35 days from 2nd dose) |  |
|  | Person-days at risk =<br>6,922,188 |  | Person-days at risk =<br>367,168 |  | Person-days at risk =<br>366,364 |  |
|  | % person<br>days at risk | # infections | % person<br>days at risk | # infections | % person<br>days at risk | # infections |
| Female | 44.30% | 6,395 | 48.5% | 411 | 48.3% | 287 |
| Male | 55.70% | 5,659 | 51.5% | 411 | 51.7% | 315 |
| General<br>Jewish | 63.90% | 10,048 | 86.7% | 743 | 94.9% | 576 |
| Ultra-Orthodox | 13.40% | 1,328 | 6.6% | 69 | 3.2% | 21 |
| Arabs | 22.80% | 678 | 6.7% | 10 | 1.8% | 5 |

**Table S2.** Characteristics of the 12-15 age group study cohorts used in the analysis. The table presents the proportion of person-days at risk instead of the proportion of individuals. Values are presented for the study period - December 26, 2021 to January 8, 2022.

| Ages 12-15<br>3rd dose effect |  |  |  |  |  |  |  |  |  |  |  |  |
| --- | --- | --- | --- | --- | --- | --- | --- | --- | --- | --- | --- | --- |
| Group | Unvaccinated* |  | 2nd dose<br>(14-60 days from 2nd dose) |  | 2nd dose<br>(60-120 days from 2nd dose) |  | 2nd dose<br>(120+ days from 2nd dose) |  | 3rd dose internal control<br>(3-7 days from 3rd dose) |  | 3rd dose<br>(14-60 days from 3rd dose) |  |
|  | Person-days at risk =<br>1,397,210 |  | Person-days at risk =<br>231,950 |  | Person-days at risk =<br>1,417,282 |  | Person-days at risk =<br>2,364,056 |  | Person-days at risk =<br>190,139 |  | Person-days at risk =<br>178,780 |  |
|  | % person<br>days at risk | #<br>infections | % person<br>days at risk | #<br>infections | % person<br>days at risk | #<br>infections | % person<br>days at risk | #<br>infections | % person<br>days at risk | #<br>infections | % person<br>days at risk | #<br>infections |
| Female | 44.50% | 2,023 | 51.00% | 102 | 50.40% | 1,240 | 49.10% | 3,616 | 48.4% | 265 | 48.9% | 99 |
| Male | 55.50% | 1,435 | 49.00% | 80 | 49.60% | 1,086 | 50.90% | 3,019 | 51.6% | 253 | 51.1% | 80 |
| General Jewish | 59.70% | 2,684 | 49.70% | 153 | 57.50% | 1,999 | 84.70% | 5,983 | 94.7% | 494 | 95.8% | 166 |
| Ultra-Orthodox | 19.60% | 627 | 7.60% | 11 | 8.20% | 217 | 6.50% | 524 | 3.1% | 20 | 2.7% | 13 |
| Arabs | 20.70% | 147 | 42.70% | 18 | 34.30% | 110 | 8.80% | 128 | 2.1% | 4 | 1.5% | 0 |

## References

1. Collie S, Champion J, Moultrie H, Bekker L-G, Gray G. Effectiveness of BNT162b2 Vaccine against Omicron Variant in South Africa. N Engl J Med 2021;NEJMc2119270.

2. SARS-CoV-2 variants in analyzed sequences [Internet]. Our World Data. [cited 2022 Apr 7];Available from: https://ourworldindata.org/grapher/covid-variants-area

3. Dorabawila V, Hoefer D, Bauer UE, Bassett MT, Lutterloh E, Rosenberg ES. Effectiveness of the BNT162b2 vaccine among children 5-11 and 12-17 years in New York after the Emergence of the Omicron Variant [Internet]. 2022 [cited 2022 May 2];2022.02.25.22271454. Available from: https://www.medrxiv.org/content/10.1101/2022.02.25.22271454v1

4. Fleming-Dutra KE, Britton A, Shang N, et al. Association of Prior BNT162b2 COVID-19 Vaccination With Symptomatic SARS-CoV-2 Infection in Children and Adolescents During Omicron Predominance. JAMA [Internet] 2022 [cited 2022 May 16];Available from: https://doi.org/10.1001/jama.2022.7493

5. Fowlkes AL, Yoon SK, Lutrick K, et al. Effectiveness of 2-Dose BNT162b2 (Pfizer BioNTech) mRNA Vaccine in Preventing SARS-CoV-2 Infection Among Children Aged 5-11 Years and Adolescents Aged 12-15 Years - PROTECT Cohort, July 2021-February 2022. MMWR Morb Mortal Wkly Rep 2022;71(11):422–8.

6. Amir O, Goldberg Y, Mandel M, et al. Protection following BNT162b2 booster in adolescents substantially exceeds that of a fresh 2-dose vaccine. Nat Commun 2022;13(1):1971.

7. Rossman H, Shilo S, Meir T, Gorfine M, Shalit U, Segal E. COVID-19 dynamics after a national immunization program in Israel. Nat Med 2021;27(6):1055–61.

8. Muhsen K, Na’aminh W, Lapidot Y, et al. A nationwide analysis of population group differences in the COVID-19 epidemic in Israel, February 2020–February 2021. Lancet Reg Health - Eur 2021;7:100130.

9. Kahn R, Schrag SJ, Verani JR, Lipsitch M. Identifying and alleviating bias due to differential depletion of susceptible people in post-marketing evaluations of COVID-19 vaccines [Internet]. Epidemiology; 2021 [cited 2021 Dec 16]. Available from: http://medrxiv.org/lookup/doi/10.1101/2021.07.15.21260595

10. Bar-On YM, Goldberg Y, Mandel M, et al. Protection of BNT162b2 vaccine booster against Covid-19 in Israel. N Engl J Med 2021;385(15):1393–400.

11. Bar-On YM, Goldberg Y, Mandel M, et al. Protection against Covid-19 by BNT162b2 Booster across Age Groups. N Engl J Med 2021;NEJMoa2115926.

12. Paediatric multisystem inflammatory syndrome temporally associated with COVID-19 (PIMS) - guidance for clinicians [Internet]. RCPCH. [cited 2022 May 16];Available from: https://www.rcpch.ac.uk/resources/paediatric-multisystem-inflammatory-syndrome-temporally-associated-covid-19-pims-guidance

